# Fear of COVID-19 among nurses in mobile COVID-19 testing units in Greece

**DOI:** 10.1101/2021.07.05.21260037

**Authors:** Petros Galanis, Emmanouela Petrogianni, Irene Vraka, Olympia Konstantakopoulou, Olga Siskou, Angeliki Bilali, Daphne Kaitelidou

## Abstract

**Background:** Mobile COVID-19 testing units are used worldwide to test quickly and easily individuals for COVID-19.

**Aim:** To assess the level of fear of COVID-19 among nurses in mobile COVID-19 testing units and compare it with demographic characteristics.

**Methods:** A cross-sectional study was conducted during November and December 2020. Study population included 57 nurses working in mobile COVID-19 testing units in Attica, Greece. We collected demographic data from the nurses, i.e. gender, age, marital status, children, living status (alone or with others), clinical experience, and chronic disease. We used the fear of COVID-19 scale to measure fear of the COVID-19 pandemic.

**Results:** The mean score on the fear of COVID-19 scale was 14.3. Among nurses, 31.6% experienced elevated fear indicative of presence of anxiety symptoms, while the respective percentages for health anxiety and post-traumatic stress symptomatology were 22.8% and 17.5%. Fear of COVID-19 was not affected by demographic variables. However, fear was higher in females, nurses who had children and nurses who lived with others. Increased clinical experience was related with decreased fear.

**Conclusions:** Creating a secure work environment for nurses in these units could decrease fear of COVID-19 and increase work performance.

## Introduction

During the coronavirus disease 2019 (COVID-19) pandemic, the frontline nurses experience enormous and various mental health challenges since they represent a high risk group for the severe acute respiratory syndrome coronavirus 2 (SARS-CoV-2) infection and they work under a tremendous psychological pressure. Depression, anxiety, stress, post-traumatic stress disorder, burnout are common among nurses during the COVID-19 pandemic [1,2]. Moreover, healthcare workers experience frequent and serious adverse events due to personal protective equipment use e.g. headaches, pressure injuries, skin injuries etc. that can affect their mental health [3].

Nurses experience extreme levels of personal fear of COVID-19, even higher than other healthcare workers and general population [4]. Moreover, healthcare workers are a vulnerable group for COVID-19 due to high seroprevalence of SARS-CoV-2 antibodies among them [5]. A systematic review found that fear of becoming infected or uncertainty is the most challenging issue among frontline healthcare workers with nurses developing the greater risk [6]. These high levels of fear can be detrimental to nurses’ physical and psychological health resulting on depression, anxiety disorders, excessive use of alcohol and tobacco, anger, insomnia, post-traumatic stress disorder, and poor self-perceived health [7,8]. Fortunately, most countries adopted COVID-19 vaccination programs from January 2021 for healthcare workers. This strategy is expected to decrease fear, anxiety and distress among healthcare workers but until now there is only a moderate level of healthcare workers intention to accept novel COVID-19 vaccines [9].

Mobile COVID-19 testing units are used worldwide to test quickly and easily asymptomatic individuals or people with symptoms of COVID-19. Nurses working in mobile COVID-19 testing units are at high risk of infecting of SARS-CoV-2 since they have close contact with suspected COVID-19 cases. However, to date, the level of fear of COVID-19 among nurses working in mobile COVID-19 testing units in unknown. Recognizing the importance of this, we aimed to assess the level of fear of COVID-19 among nurses in mobile COVID-19 testing units and compare it with demographic characteristics.

## Material and method

### Study design and participants

A cross-sectional study was conducted during November and December 2020. During this time, the total number of nurses working in mobile COVID-19 testing units in Attica, Greece was 60. District of Attica is the biggest one in Greece. All nurses have been working in mobile COVID-19 testing units at least for four months. We invited all nurses to participate in our study and the response rate was 95% (57 out of 60). These nurses collected specimens from suspected COVID-19 cases to perform diagnostic test for SARS-CoV-2. Specimens were collected from individuals in several places, e.g. nursing homes, social services facilities, temporary reception facilities for refugees and immigrants, day-care centers etc. Physical copies of the questionnaire were filled anonymously by the nurses. Institutional ethical approval from the Department of Nursing, National and Kapodistrian University of Athens (reference number: 347) was obtained prior to conducting the study. Moreover, all nurses provided informed consent to participate in the study.

### Measures

We collected demographic data from the nurses, i.e. gender, age, marital status, children, living status, clinical experience, and chronic disease.

We used the fear of COVID-19 scale (FCV-19S) that evaluates fear of the COVID-19 pandemic [10]. In particular, the validated Greek language version of the FCV-19S was used [11]. The FCV-19S comprises seven items, each with a five-point Likert scale of answers from strongly disagree (1) to strongly agree (5). Total score of the FCV-19S ranges from 7 to 35 with higher values indicate greater fear. Cronbach’s alpha for fear of COVID-19 scale was 0.79 indicating good internal consistency.

We used scores on the FCV-19S as a proxy for anxiety, health anxiety, and post-traumatic stress symptomatology according to the proposed cut-off points for the Greek version of the scale [12]. The suggested cut-off points are 16.5 for anxiety, 17.5 for health anxiety, and 18.5 for post-traumatic stress symptoms. Scores above these cut-off points denote elevated fear and a higher probability of anxiety, health anxiety, and post-traumatic stress symptoms.

### Statistical analysis

Categorical variables are presented as numbers (percentages). We used mean (standard deviation) to describe the continuous variables that followed normal distribution, and median (interquartile range) to describe the continuous variables with skewness. The Kolmogorov-Smirnov test was used to test the normality of the distribution of the continuous variables. Demographic data of nurses were considered as the independent variables, while fear of COVID-19, anxiety, health anxiety, and post-traumatic stress symptomatology were considered as the dependent variables. We used the chi-square test and Fisher’s exact test to compare categorical variables. Independent samples t-test and Mann-Whitney test were used to compare continuous data across groups in case of normal or skewed distribution respectively. Correlation between continuous data was assessed with Spearman’s correlation coefficient. All tests of statistical significance were two-tailed, and p-values < 0.05 were considered significant. Statistical analysis was performed with IBM SPSS Version 21.0.

## Results

Study population included 57 nurses with a mean age of 33.3 years (standard deviation, 8.7). The majority of nurses was females (89.5%) and singles (50.9%). Among nurses, 21.1% had children, 80.7% lived with others and 12.3% have suffered from a chronic disease. Mean years of clinical experience were 6.9 (standard deviation, 8.1).

The mean score on the fear of COVID-19 scale was 14.3 (standard deviation, 8.2). The mean scores regarding the fear of COVID-19 scale and differences based on demographic data are shown in Table 1. No statistically significant differences were found. Fear of COVID-19 was higher among females, married, nurses living with others, and nurses without a chronic disease. Increased clinical experience was related with decreased fear.

**Table 1.** Item wise and total mean score comparison on fear of COVID-19 scale based on demographic characteristics.

| Demographic characteristics | Item |  |  |  |  |  |  |  |
| --- | --- | --- | --- | --- | --- | --- | --- | --- |
|  | I am most afraid of COVID-19 | It makes me uncomfortable to think about COVID-19 | My hands become clammy when I think about COVID-19 | I am afraid of losing my life because of COVID-19 | When watching news and stories about COVID-19 on social media, I become nervous or anxious | I cannot sleep because I'm worrying about getting COVID-19 | My heart races or palpitates when I think about getting COVID-19 | Total scale |
| Total sample | 2.72 (1.03) | 2.61 (1.15) | 1.63 (0.79) | 1.79 (0.98) | 2.63 (1.13) | 1.39 (0.68) | 1.53 (0.76) | 14.30 (4.43) |
| Gender |  |  |  |  |  |  |  |  |
| Females | 2.75 (1.02) | 2.69 (1.09) | 1.65 (0.79) | 1.78 (0.95) | 2.69 (1.12) | 1.41 (0.70) | 1.51 (0.70) | 14.47 (4.30) |
| Males | 2.50 (1.23) | 2.00 (1.55) | 1.50 (0.84) | 1.83 (1.33) | 2.17 (1.17) | 1.17 (0.41) | 1.67 (1.21) | 12.83 (5.67) |
| p-value<sup>a</sup> | 0.59 | 0.17 | 0.67 | 0.91 | 0.29 | 0.41 | 0.64 | 0.40 |
| Age (years)<sup>b</sup> | -0.06 | -0.04 | 0.05 | -0.04 | -0.03 | 0.06 | 0.02 | -0.03 |
| p-value<sup>b</sup> | 0.64 | 0.78 | 0.70 | 0.75 | 0.84 | 0.66 | 0.87 | 0.85 |
| Marital status |  |  |  |  |  |  |  |  |
| Single/divorced | 2.57 (0.88) | 2.60 (1.06) | 1.63 (0.84) | 1.83 (0.89) | 2.63 (1.11) | 1.37 (0.69) | 1.51 (0.70) | 14.14 (3.69) |
| Married | 2.95 (1.21) | 2.64 (1.29) | 1.64 (0.73) | 1.73 (1.12) | 2.64 (1.18) | 1.41 (0.67) | 1.55 (0.86) | 14.55 (5.48) |
| p-value<sup>a</sup> | 0.17 | 0.91 | 0.97 | 0.71 | 0.98 | 0.84 | 0.88 | 0.74 |
| Children |  |  |  |  |  |  |  |  |
| Yes | 2.58 (1.00) | 2.67 (1.16) | 1.75 (0.62) | 1.83 (0.94) | 2.25 (1.14) | 1.67 (0.78) | 1.75 (0.75) | 14.50 (5.63) |
| No | 2.76 (1.05) | 2.60 (1.16) | 1.60 (0.84) | 1.78 (1.00) | 2.73 (1.12) | 1.31 (0.63) | 1.47 (0.76) | 14.24 (4.12) |
| p-value<sup>a</sup> | 0.61 | 0.86 | 0.57 | 0.86 | 0.19 | 0.11 | 0.25 | 0.86 |
| Living |  |  |  |  |  |  |  |  |
| Alone | 2.36 (0.67) | 2.18 (0.87) | 1.64 (0.81) | 1.45 (0.69) | 2.64 (1.03) | 1.18 (0.41) | 1.18 (0.41) | 12.63 (3.26) |
| With others | 2.80 (1.09) | 2.72 (1.19) | 1.63 (0.80) | 1.87 (1.02) | 2.63 (1.16) | 1.43 (0.72) | 1.61 (0.80) | 14.70 (4.60) |
| p-value<sup>a</sup> | 0.21 | 0.17 | 0.98 | 0.21 | 0.99 | 0.27 | 0.1 | 0.10 |
| Clinical experience (years)<sup>b</sup> | -0.15 | -0.15 | 0.03 | -0.14 | 0.06 | 0.10 | -0.06 | -0.11 |
| p-value<sup>b</sup> | 0.27 | 0.27 | 0.83 | 0.29 | 0.68 | 0.46 | 0.6 | 0.42 |
| Chronic disease |  |  |  |  |  |  |  |  |
| Yes | 2.71 (0.95) | 2.29 (1.11) | 1.29 (0.49) | 1.43 (0.79) | 2.86 (0.90) | 1.43 (0.79) | 1.43 (0.79) | 13.43 (4.83) |
| No | 2.72 (1.05) | 2.66 (1.15) | 1.68 (0.82) | 1.84 (1.00) | 2.60 (1.16) | 1.38 (0.67) | 1.54 (0.76) | 14.42 (4.41) |
| p-value<sup>a</sup> | 0.99 | 0.42 | 0.10 | 0.30 | 0.58 | 0.86 | 0.72 | 0.58 |

Values are expressed as mean (standard deviation) unless otherwise is indicated
<sup>a</sup> independent samples t-test
<sup>b</sup> Spearman’s correlation coefficient

We used the suggested cut-off points for anxiety (16.5), health anxiety (17.5), and post-traumatic stress symptomatology (18.5) to classify the study population (Table 2). Among nurses, 31.6% experienced elevated fear indicative of presence of anxiety symptoms, while the respective percentages for health anxiety and post-traumatic stress symptomatology were 22.8% and 17.5%. We did not find statistically significant differences between demographic data and anxiety, health anxiety, and post-traumatic stress symptomatology. Females, married, nurses with children and those living with others reported higher fear levels and thus higher levels of anxiety, health anxiety, and post-traumatic stress symptoms. Also, older nurses displayed higher levels of anxiety, health anxiety, and post-traumatic stress symptoms.

**Table 2.** Study population classification according to cut-off points (i.e. 16.5 for anxiety, 17.5 for health anxiety, and 18.5 for post-traumatic stress symptomatology).

| Demographic characteristics | Anxiety |  | P-value | Health anxiety |  | P-value | Post-traumatic stress symptomatology |  | P-value |
| --- | --- | --- | --- | --- | --- | --- | --- | --- | --- |
|  | Normal fear | Elevated fear |  | Normal fear | Elevated fear |  | Normal fear | Elevated fear |  |
| Total sample | 39 (68.4) | 18 (31.6) |  | 44 (77.2) | 13 (22.8) |  | 47 (82.5) | 10 (17.5) |  |
| Gender |  |  | 0.92<sup>a</sup> |  |  | 0.52<sup>a</sup> |  |  | 0.28<sup>a</sup> |
| Females | 4 (66.7) | 2 (33.3) |  | 4 (66.7) | 2 (33.3) |  | 4 (66.7) | 2 (33.3) |  |
| Males | 35 (68.6) | 16 (31.4) |  | 40 (78.4) | 11 (21.6) |  | 43 (84.3) | 8 (15.7) |  |
| Age (years)<sup>b</sup> | 32.59 (8.37) | 34.83 (9.32) | 0.37<sup>c</sup> | 32.61 (8.72) | 35.62 (8.37) | 0.28<sup>c</sup> | 33.02 (8.89) | 34.60 (7.99) | 0.61<sup>c</sup> |
| Marital status |  |  | 0.07<sup>a</sup> |  |  | 0.19<sup>a</sup> |  |  | 0.42<sup>a</sup> |
| Single/divorced | 27 (77.1) | 8 (22.9) |  | 29 (82.9) | 6 (17.1) |  | 30 (85.7) | 5 (14.3) |  |
| Married | 12 (54.5) | 10 (45.5) |  | 15 (68.2) | 7 (31.8) |  | 17 (77.3) | 5 (22.7) |  |
| Children |  |  | 0.17<sup>d</sup> |  |  | 0.44<sup>d</sup> |  |  | 0.42<sup>d</sup> |
| Yes | 6 (50.0) | 6 (50.0) |  | 8 (66.7) | 4 (33.3) |  | 9 (75.0) | 3 (25.0) |  |
| No | 33 (73.3) | 12 (26.7) |  | 36 (80.0) | 9 (20.0) |  | 38 (84.4) | 7 (15.6) |  |
| Living |  |  | 0.15<sup>d</sup> |  |  | 0.43<sup>d</sup> |  |  | 0.68<sup>d</sup> |
| Alone | 10 (90.9) | 1 (9.1) |  | 10 (90.9) | 1 (9.1) |  | 10 (90.9) | 1 (9.1) |  |
| With others | 29 (63.0) | 17 (37.0) |  | 34 (73.9) | 12 (26.1) |  | 37 (80.4) | 9 (19.6) |  |
| Clinical experience (years)<sup>e</sup> | 4.0 (10.0) | 2.5 (11.0) | 0.48<sup>f</sup> | 3.0 (10.0) | 5.0 (12.0) | 0.96<sup>f</sup> | 3.0 (10.0) | 3.5 (11.0) | 0.75<sup>f</sup> |
| Chronic disease |  |  | 0.41<sup>d</sup> |  |  | 0.99<sup>d</sup> |  |  | 0.99<sup>d</sup> |
| Yes | 6 (85.7) | 1 (14.3) |  | 6 (85.7) | 1 (14.3) |  | 6 (85.7) | 1 (14.3) |  |
| No | 33 (66.0) | 17 (34.0) |  | 38 (76.0) | 12 (24.0) |  | 41 (82.0) | 9 (18.0) |  |

Values are expressed as number (percentage) unless otherwise is indicated
<sup>a</sup> chi-square test
<sup>b</sup> mean (standard deviation)
<sup>c</sup> independent samples t-test
<sup>d</sup> Fisher's exact test
<sup>e</sup> median (interquartile range)
<sup>f</sup> Mann-Whitney test

## Discussion

To our knowledge, this is the first study worldwide that determined the levels of fear of COVID-19 among nurses in mobile COVID-19 testing units. In the current study, the mean score on the fear of COVID-19 scale was 14.3. To date, three studies in Mexico, Philippines and Turkey have used the fear of COVID-19 scale to measure frontline nurses’ fear finding mean values higher than our study (from 18.2 to 19.3) [4,13,14]. Interestingly, nurses in our study had less fear than the general population where the mean scale score for the fear of COVID-19 measure ranges from 16.6 to 19.4 [15]. In general, frontline nurses are under extreme psychological pressure and suffer from fear of contracting SARS-CoV-2 and infecting patients, colleagues, and family members [16]. On the other hand, nurses in mobile COVID-19 testing units are equipped with the appropriate personal protective equipment and they are well trained to use this equipment. This scenario could probably reduces nurses’ fear for COVID-19 since personal protective equipment shortages increase anxiety, depression, burnout and stress [1]. Also, availability of personal protective equipment makes nurses feel self-confident creating a safe work environment for nurses, their colleagues and patients [17].

Applying the suggested cut-off points [12] for the fear of COVID-19 scale, we found that a considerable percentage of nurses in our study developed psychological symptoms. In particular, 31.6%, 22.8% and 17.5% of nurses had anxiety, health anxiety, and post-traumatic stress symptomatology respectively. Literature confirms this finding since mental and psychological problems are common among nurses during the COVID-19 pandemic, e.g. depression, anxiety, stress, sleep disturbances, and burnout [2]. Moreover, nurses suffer higher rates of COVID-19 fear than other healthcare workers since nurses are in close contact with COVID-19 patients for long periods of time [4,13].

Fear of COVID-19 was not affected by demographic variables in our study. However, it is worth of note that fear was higher in females, nurses who had children and nurses who lived with others. Several studies confirm that female nurses have more fear and show stronger self-perceived threat from the COVID-19 [13,18]. Previous studies have suggested that nurses who have children experience higher levels of fear since they are afraid of getting infected with the SARS-CoV-2 and transmitting it to close family members [18].

The nature of our study was exploratory and thus had several limitations. First, the sample size was relatively small and our study was performed within one district of Greece. However, all the nurses working in mobile COVID-19 testing units in this district were invited to participate and the response rate was very high. Next, we investigated only a few demographic variables as possible determinants of fear of COVID-19 and further studies should explore other personal and psychological variables. Additionally, we used a standardized and valid instrument to measure fear of COVID-19 but an information bias is still probable. Another limitation is that our study was conducted during November and December 2020 and further research should be carried out since fear of COVID-19 is an evolving issue.

Despite these limitations, the results of our study could provide useful information since mobile COVID-19 testing units are used worldwide as another weapon against the COVID-pandemic. Creating a secure work environment for nurses in these units could decrease fear of COVID-19 and increase work performance.

## Data Availability

Data will be available after reasonable request

## References

1. Sampaio F, Sequeira C, Teixeira L. Nurses’ Mental Health During the Covid-19 Outbreak: A Cross-Sectional Study. J Occup Environ Med. 2020;62:783–7.

2. Galanis P, Vraka I, Fragkou D, Bilali A, Kaitelidou D. Nurses’ burnout and associated risk factors during the COVIDLJ19 pandemic: A systematic review and metaLJanalysis. J Adv Nurs. 2021; under press.

3. Galanis P, Vraka I, Fragkou D, Bilali A, Kaitelidou D. Impact of personal protective equipment use on health care workers’ physical health during the COVID-19 pandemic: A systematic review and meta-analysis. Am J Infect Control. 2021; under press.

4. Saracoglu KT, Simsek T, Kahraman S, Bombaci E, Sezen Ö, Saracoglu A, et al. The Psychological Impact of COVID-19 Disease is more Severe on Intensive Care Unit Healthcare Providers: A Cross-sectional Study. Clin Psychopharmacol Neurosci. 2020;18:607–15.

5. Galanis P, Vraka I, Fragkou D, Bilali A, Kaitelidou D. Seroprevalence of SARS-CoV-2 antibodies and associated factors in healthcare workers: a systematic review and meta-analysis. J Hosp Infect. 2021;108:120–34.

6. Cabarkapa S, Nadjidai SE, Murgier J, Ng CH. The psychological impact of COVID-19 and other viral epidemics on frontline healthcare workers and ways to address it: A rapid systematic review. Brain Behav Immun. 2020;8:100144.

7. Garfin DR, Silver RC, Holman EA. The novel coronavirus (COVID-2019) outbreak: Amplification of public health consequences by media exposure. Health Psychol. 2020;39:355–7.

8. Sloan M, Haner M, Graham A, Cullen FT, Pickett J, Jonson CL. Pandemic Emotions: The Extent, Correlates, and Mental Health Consequences of Personal and Altruistic Fear of COVID-19 [Internet]. SocArXiv; 2020 May. Available from: https://osf.io/txqb6

9. Galanis PA, Vraka I, Fragkou D, Bilali A, Kaitelidou D. Intention of health care workers to accept COVID-19 vaccination and related factors: a systematic review and meta-analysis [Internet]. Public and Global Health; 2020 Dec. Available from: http://medrxiv.org/lookup/doi/10.1101/2020.12.08.20246041

10. Ahorsu DK, Lin C-Y, Imani V, Saffari M, Griffiths MD, Pakpour AH. The Fear of COVID-19 Scale: Development and Initial Validation. Int J Ment Health Addict. 2020;1– 9.

11. Tsipropoulou V, Nikopoulou VA, Holeva V, Nasika Z, Diakogiannis I, Sakka S, et al. Psychometric Properties of the Greek Version of FCV-19S. Int J Ment Health Addict. 2020:1–10.

12. Nikopoulou VA, Holeva V, Parlapani E, Karamouzi P, Voitsidis P, Porfyri GN, et al. Mental Health Screening for COVID-19: a Proposed Cutoff Score for the Greek Version of the Fear of COVID-19 Scale (FCV-19S). Int J Ment Health Addict. 2020;1–14.

13. García-Reyna B, Castillo-García GD, Barbosa-Camacho FJ, Cervantes-Cardona GA, Cervantes-Pérez E, Torres-Mendoza BM, et al. Fear of COVID-19 Scale for Hospital Staff in Regional Hospitals in Mexico: a Brief Report. Int J Ment Health Addict. 2020;1– 12.

14. Labrague LJ, Santos JAA. Fear of COVIDLJ19, psychological distress, work satisfaction and turnover intention among frontline nurses. J Nurs Manag. 2020; under press.

15. Gritsenko V, Skugarevsky O, Konstantinov V, Khamenka N, Marinova T, Reznik A, et al. COVID 19 Fear, Stress, Anxiety, and Substance Use Among Russian and Belarusian University Students. Int J Ment Health Addict. 2020;1–7.

16. Galehdar N, Kamran A, Toulabi T, Heydari H. Exploring nurses’ experiences of psychological distress during care of patients with COVID-19: a qualitative study. BMC Psychiatry. 2020;20:489.

17. Sharma A, Gupta P, Jha R. COVID-19: Impact on Health Supply Chain and Lessons to Be Learnt. J Health Manag. 2020;22:248–61.

18. Sakib N, Akter T, Zohra F, Bhuiyan AKMI, Mamun MA, Griffiths MD. Fear of COVID-19 and Depression: A Comparative Study Among the General Population and Healthcare Professionals During COVID-19 Pandemic Crisis in Bangladesh. Int J Ment Health Addict. 2021;1–17.

